# Predictors of voluntary uptake of modern contraceptive methods in rural Sindh, Pakistan

**DOI:** 10.1101/2023.09.04.23295034

**Authors:** Zahid Memon, Wardah Ahmed, Abeer Mian, Muhammad Jawwad, Shah Muhammad, Abdul Qayyum Noorani, Zulfiqar Bhutta, Hora Soltani

## Abstract

**Introduction:** The use of modern contraceptive methods (MCMs) in Pakistan has been stagnant for the last decade. In Sindh, current contraceptive use is at 28.9%, of which 25% is MCMs use. Such a low uptake translates into high unmet need 17% amongst married women. To bridge the gap between the health system and beneficiaries, there is a need to assess predictors that influence voluntary uptake of MCMs among women, at the health services and individual levels.

**Methods:** A cross-sectional household survey was conducted in two districts of Sindh, Pakistan namely Matiari and Badin. In total, 1684 Married Women of Reproductive Age (MWRA) 15-49 years were interviewed. For the selection of eligible respondents, a two-stage stratified cluster sampling strategy was used. Univariate and multivariable logistic regression was used to determine the predictive factors for the increase in the use of MCM.

**Results:** Mean age was 32.3 ±SD 7.1 years. Average number of children per woman was 4.0 ± 2.0. Use of modern methods of contraceptive was 26.1% [n=441).Statistically significant socio demographic predictors of MCM included: Number of children 4 or more (AOR 5.234 95%CI 2.78-9.84), Mother having primary education (AOR 1.730 95% CI 1.26-2.36), and Husband having middle education [AOR 1.69 95% CI 1.03 – 2.76).Maternal health services indicators included postnatal checkup of mother (AOR 1.46 95% CI 1.09 – 2.05); women who were visited by Lady Health Workers in their postnatal period and were counseled on family planning (AOR 1.83 95% CI 1.386 - 2.424).

**Conclusion:** Voluntary uptake of modern contraceptive methods is higher in women having 2 or more children, having primary education and husband having middle education. Significantly, receiving post-natal checkup at facility, and Lady Health Worker visit after delivery have more likelihood to opt for contraception. Additionally, young couple counseling on family planning is imperative to bridge the gap between knowledge and its translation into practice. There is also a need to focus on the provision of integrated family planning and maternal, newborn, and child health services through facility-based and community engagement platforms.

## Introduction

According to the 7^th^ population and housing census 2023, the population of Pakistan stood at 241.5 million, with an annual growth rate of 2.5% making it 5^th^ populous country in the world [1]. This growth rate is double compared to South Asia’s average rate (1.2%). It is forecasted to reach 403 million inhabitants in 2050 [2]. This rapid and for the most part, uncontrolled population growth has contributed to the economic and social crisis. Pakistan faces and affects the health of women and children disproportionately. Overall, 63% of the population lives in rural areas with low accessibility to health care services.

Global evidence indicates that such rampant population growth rates are also associated with poor coverage of family planning programs and unmet needs for contraception. Investments in social determinants and programs for supporting and scaling up modern contraception rates play a key role in reducing maternal and infant mortality and improving human, social, and economic development on an individual and national level [3, 4, 5].

Despite the benefits of family planning, the use of modern contraceptive methods (MCMs) in Pakistan has been stagnant for the last decade. The country-wide contraceptive prevalence rate (CPR) stands at 34%, with MCMs comprising 26% of this rate [6]. Within Pakistan, the second largest province of Sindh reports contraceptive use at 28.9%, of which 25% is attributed to MCM use. Such low uptake rates translate into high unmet need 17% amongst married women who are fecund and desire birth spacing or limit the number of children they have. Despite these needs, they are currently not accessing or utilizing any methods of contraception. [7].

The reasons for this low MCM access and utilization are varied and multifaceted. A major contributing factor is the prevalence of early marriages, which constitutes 16.3% of all marriages and primarily involves those between the ages 15 and 19 years [7].

In keeping with the need to improve these rates, previous studies have attempted to explore various predictors of modern contraceptive uptake. Predictors of uptake covered in the local literature include a woman’s age, education, wealth status, and positive attitude towards contraception at an individual level [8]. Further studies have reported that interventions encompassing elements of social franchising, mid-level health care providers (community midwives) engagement, and mass media participation substantially increase the prevalence of contraceptive use [9, 10]. Thus, these determinants provide an important evidence base to effectively design, implement and further evaluate interventions aimed at increasing contraceptive uptake.

Addressing these determinants requires a distinct strategy, intervention, or program. However, introducing numerous new programs external to the existing healthcare infrastructure is unfeasible in light of the country’s economic instability combined with a lack of political will and a heavily underfunded healthcare system [11]. Therefore, the main aim of this paper is to assess the predictors associated with the uptake of MCMs in the domains of sociodemographic, reproductive, and maternal health with corresponding knowledge, attitude, and use amongst married women of reproductive age (MWRA) between 15-49 years in two districts of rural Sindh. Protocol paper has been published for more details on methodology [12].

## Methods and Materials

This was a cross-sectional study with face-to-face interviews of married women of reproductive age at household level, using a structured questionnaire, as the pregnancies in Pakistan primarily occur following marriage [13]. This survey was conducted in two districts of the province of Sindh: Matiari and Badin (Figure 1).

**Figure 1.**
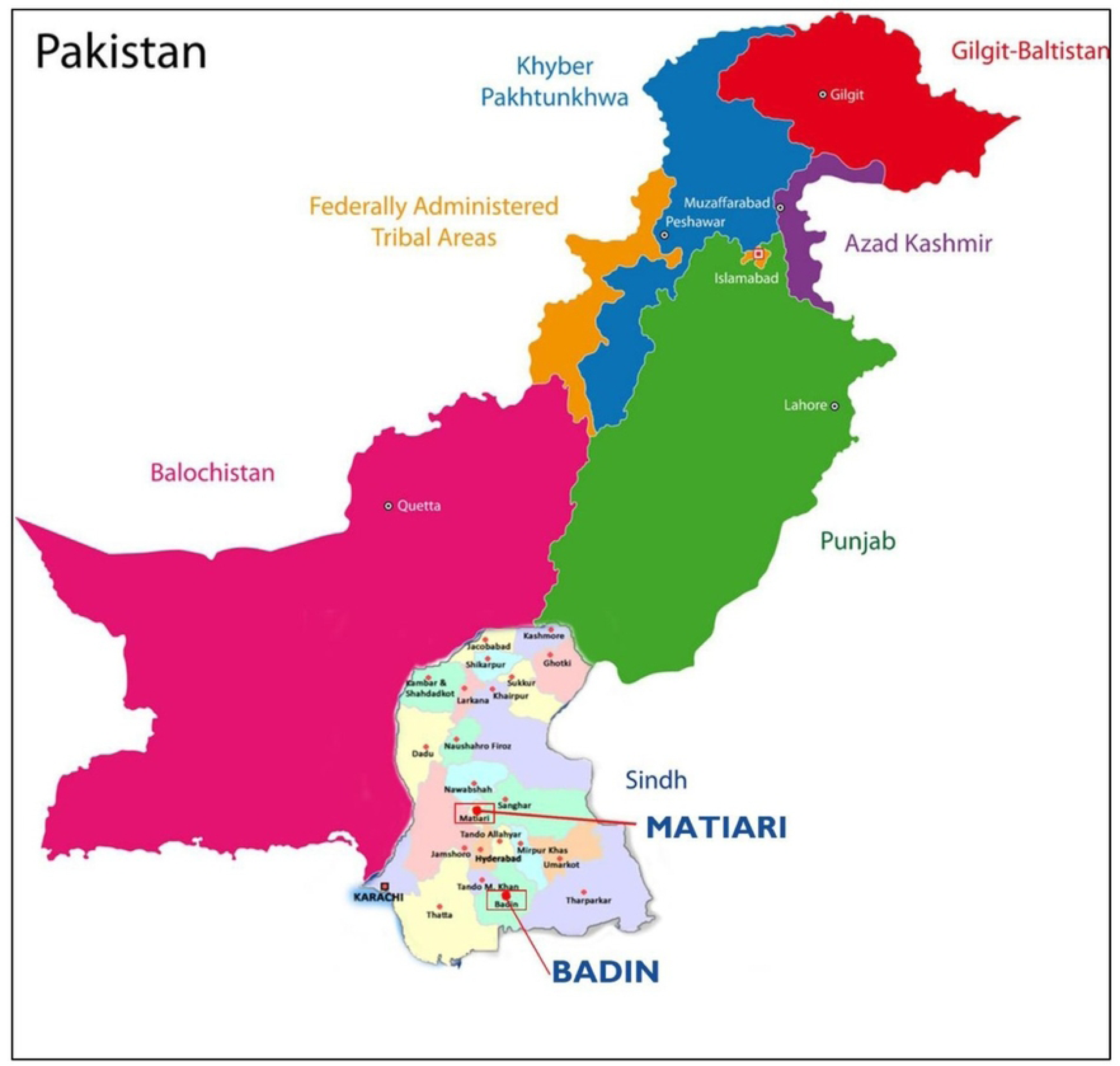
Study Sites – Badin and Matiari Districts of Province Sindh, Pakistan. The selection of districts was based on propensity matching score further details could be obtained from protocol paper [12]. Data was collected on all relevant program indicators, including the study population’s modern contraceptive prevalence rate (mCPR), antenatal and postnatal care services, skilled birth attendance, and socio-economic and demographic status.

### Sample size estimation

As per the Multiple Indicator Cluster Survey for Sindh 2018-2019 [7], the CPR reported for Sindh province was 28.9%. Based on the CPR, with 95% CI and 80% power, an assumed design effect of 1.5 and a 7% non-response rate, a sample size of 880 MWRA (15-49 years) per district was estimated.

The targeted population for this study consisted of MWRA between 15-49 years of age residing in the study districts within the catchment population of public sector health facilities. The exclusion criteria for the sample included: (i) families who had moved into the project area within three months with no intent to stay for a prolonged period in the project area and (ii) women of reproductive age with mental disorders.

A two-stage stratified cluster sampling strategy was used to select eligible respondents. Clusters were created with at least 100-150 households and were established as primary sampling units (PSUs). In the first sampling stage, clusters were randomly selected from each district’s list of clusters (sampling frame). Subsequently, a line listing operation was conducted to determine eligible households assigned as secondary sampling units (SSUs). In the second stage, 20 eligible households with at least one MWRA were selected using systematic sampling to reach the calculated sample size (covering 44 clusters in each district).

### Survey questionnaire

The household questionnaire was adopted from the Pakistan Demographic and Health Survey (PDHS) questionnaire for women of reproductive age [6]. The questionnaire was then pre-tested in 5% of the total sample population, then translated into the local Sindhi language. A customized digital application and software were developed for digital data collection.

### Survey training

Before data collection, a five-day training workshop on the data collection process was conducted for field teams (directly involved in field-based data collection). All sections of the questionnaires were discussed in detail, followed by a mock field-based data collection activity for data collectors supervised by the senior research staff. In each district, there are three teams consisting of four senior research assistants and one research associate who have received 16 years of education in social or medical allied sciences. The teams were carefully formed based on educational qualifications and job descriptions to effectively conduct the survey. The last training session was dedicated to feedback and troubleshooting based on issues reported from the mock exercise. The sessions involved extensive training on informed consent, manual and digital data collection processes. Training manuals were developed and distributed to the data collectors and questionnaires for pilot testing in the field.

### Data collection and data confidentiality

The study was conducted in each district by the assigned data collection team consisting of one team leader and four data collectors. Before the data collection process commenced, line listing was undertaken in selected clusters. Verbal informed consent was taken from married respondents and details of the risks and benefits of their participation were explained in the presence of an impartial witness and guardian if respondent was under 18 years of age. They were given an opportunity to ask any questions they had regarding the study, after which their consent was obtained (except for those refusing to participate).

Data was collected between 1^st^ October 2020 and 31^st^ December 2020, on personal digital assistants (Model Samsung SM T289) through an Android application designed for field-based data collection. Data collection duration was 45-60 minutes. All data was uploaded electronically, analyzed, and displayed on the virtual project dashboard. To maintain confidentiality, coded data was stored in a password-protected and encrypted system, ensuring that only authorized individuals had access to it.

### Data analysis

The collected data was then analyzed in STATA version 17 (Stata Corp, Texas). Descriptive statistics, bivariate statistical tests, chi-square tests, and independent sample t-tests were used to summarize the data. Categorical variables were presented in frequency and percentages, while mean and standard deviation were used for continuous variables. Furthermore, univariate and multivariable logistic regression was used to determine the predictive factors associated with increase in the use of modern methods of contraceptives amongst married women of reproductive age. Variables significant at a p-value of less than 0.20 in the univariate analysis were included in multivariable logistic regression to measure association at 95% confidence interval [14]. The final model was constructed using backward elimination; variables were retained if the p-value was less than 0.05 [15].

### Ethical approval

Ethical approval was received from the Ethical Review Committee (ERC) of the Aga Khan University on July 16, 2020 (2020-3606-18261). The study protocol was approved by the National Bioethics Committee, Pakistan (4-87/NBC-514/22/857).

## Results

From the total sample size of 1,760, 96% (n=1,684) of MWRA were interviewed. There was a high response rate of 96%, as projected in the interview-based studies at the household level.

### Sociodemographic characteristics of MWRA

In terms of sociodemographic characteristics in table 1, the mean age of the study participants was 32.3years, with a standard deviation (SD) of 7.1 years. Among respondents, 27.0% (n=453) having formal education. The average number of children per woman was 4.0± 2.0, and the average number of members living in the household was 7.2 ± 2.7.

**Table 1:** Socio demographic characteristics of MWRA(n=1,684)

| Variable | Overall |  |
| --- | --- | --- |
|  | mean | SD |
| **Household Size** | 7.2 | ±2.7 |
| **Respondent's age** | 32.3 | ± 7.1 |
| **Husband's Age** | 36.4 | ± 8.5 |
| **Number of Children (Parity)** | 4.0 | ±2.0 |
| **Number of Pregnancies** | 4.2 | ±2.3 |
|  | **n** | **%** |
| **Mother's Education** |  |  |
| No education | 1231 | 73.0 |
| Primary | 224 | 13.3 |
| Middle | 58 | 3.4 |
| Secondary | 68 | 4.0 |
| Intermediate or above | 103 | 6.1 |
| **Wealth Quantile** |  |  |
| Poorest | 335 | 19.8 |
| Poor | 336 | 19.9 |
| Middle | 339 | 20.1 |
| Rich | 338 | 20.0 |
| Richest | 336 | 19.9 |

### Current use of contraceptives

Table 2 showed that from a total of 1,684 married women, 76.2% (n=1255) reported that they heard at least one method of family planning to delay pregnancy. The difference in knowledge of contraceptives was found to be significant in current users compared to non-users. Almost half of the women who reported having knowledge of contraceptives had used family planning methods in the past. Ever use of any contraceptive method to delay or avoid pregnancy was observed to be significant amongst respondents.

**Table 2:** Current use of contraceptives reported by MWRA (n=1,684)

| Characteristics | Overall |  |
| --- | --- | --- |
|  | n | % |
| **Ever heard of any method to delay pregnancy** | 1255 | 76.2 |
| **Ever used any method to delay/avoid pregnancy** | 625 | 38.5 |
| **Current use of contraceptive** | 463 | 27.4 |
| **Any modern method** | 441 | 26.1 |
| Pills | 83 | 5.0 |
| Injectables | 63 | 3.7 |
| Condoms | 87 | 5.1 |
| Intra Uterine Devices(IUD) | 12 | 0.7 |
| Implant | 41 | 2.4 |
| Standard Days Method (SDM) | 1 | 0.1 |
| Lactational Amenorrhea Method (LAM) | 6 | 0.3 |
| Female sterilization | 109 | 6.5 |
| Male sterilization | 1 | 0.1 |
| **Traditional method** | 22 | 1.3 |
| Rhythm method | 3 | 0.1 |
| Withdrawal | 19 | 1.1 |

The contraceptive prevalence rate was 27.4 % (n=462) in the sampled population. Furthermore, the overall use of modern contraceptive methods was observed to be slightly lower at 26.1% (n=441). Among different methods, the most common methods reported by the respondents were condoms (n=87; 5.1%) followed by pills (n=83; 5.0%) and injectables (n=63; 3.7%). Out of all the long-acting reversible methods, less than two percent of the sample population reported using implants (n=41; 2.4%) and IUDs (n=12; 0.7%). The use of implants was seen as preferential to the use of IUDs across both districts.

Moreover, the lactation amenorrhea method (LAM) use was reported by 6 women. However, one woman reported the use of standard days methods (SDM). Moreover, among permanent methods of contraception, 6.5% (n=109) of the respondents reported female sterilization and only one respondent reported male sterilization method use.

The prevalence of traditional methods was less than two percent amongst all surveyed women (n=22; 1.3%). Among such practices, 1.1% (n=19) of observations were recorded for the withdrawal method, which was found to be the most common, followed by the rhythm method (n=3; 0.1%). Additionally, distribution of modern contraceptive methods among socio demographic characteristics attached as supporting information (S1_File).

### Predictors for use of modern contraceptive methods (MCM)

Table 3 shows the univariate and multivariate analysis of the predictors associated with the use of modern contraceptives.

**Table 3:** Predictors for use of modern contraceptive method (MCM) among Married Women of Reproductive age (MWRA-15-49years)

| Variable | Modern Contraceptive Use |  | Univariate Analysis |  | Multivariate Analysis |  |
| --- | --- | --- | --- | --- | --- | --- |
|  | Yes | No | Crude Odd Ratio | 95% CI | Adjusted Odds Ratio | 95% CI |
| **Number of children** |  |  |  |  |  |  |
| 1 | 30 (6.8%) | 230 (18.7%) | 1 | 1 | 1 | 1 |
| 2 | 62 (14.3%) | 210 (16.4%) | 2.395* | 1.425 – 4.024 | 2.641* | 1.445 - 4.825 |
| 3 | 60 (12.4%) | 203 (15.9%) | 2.140* | 1.284 – 3.567 | 2.644* | 1.401 - 4.990 |
| 4 or more children | 289 (66.6%) | 600 (49.0%) | 3.739* | 2.274 - 6.149 | 5.234* | 2.784 - 9.842 |
| **Mother's age [in years]** |  |  |  |  |  |  |
| 15-19 | 7 (2.2%) | 44 (3.9%) | 1 | 1 |  |  |
| 20-24 | 35 (8.1%) | 143 (12.0%) | 1.186 | 0.562 - 2.505 | - | - |
| 25-29 | 58 (12.7%) | 257 (19.1%) | 1.171 | 0.551 - 2.487 | - | - |
| 30-34 | 117 (24.9%) | 310 (25.0%) | 1.747 | 0.806 - 3.787 | - | - |
| 35-39 | 119 (28.5%) | 245 (19.6%) | 2.556* | 1.345 - 4.857 | - | - |
| 40 or above | 105 (23.7%) | 244 (20.5%) | 2.025* | 0.983 - 4.170 | - | - |
| **Mother's education** |  |  |  |  |  |  |
| No education | 289 (67.3%) | 942 (76.0%) | 1 | 1 | 1 | 1 |
| Primary | 79 (17.4%) | 145 (12.3%) | 1.599* | 1.211 - 2.112 | 1.730* | 1.264 - 2.367 |
| Middle | 20 (4.4%) | 38 (3.0%) | 1.629* | 0.970 – 2.737 | 1.671 | 0.950 - 2.940 |
| Secondary | 20 (4.2%) | 48 (3.8%) | 1.240 | 0.716 - 2.150 | 1.534 | 0.820 - 2.867 |
| Intermediate or above | 33 (6.7%) | 70 (4.8%) | 1.579 | 0.931 - 2.677 | 1.819 | 1.017 - 3.254 |
| **Husband's education** |  |  |  |  |  |  |
| No education | 158 (38.0%) | 501 (39.9%) | 1 | 1 | 1 | 1 |
| Primary | 72 (15.6%) | 253 (21.1%) | 0.777 | 0.527 - 1.146 | 0.713 | 0.481 - 1.055 |
| Middle | 42 (8.9%) | 67 (5.0%) | 1.879* | 1.201 – 2.939 | 1.695* | 1.039 – 2.762 |
| Secondary | 61 (12.8%) | 152 (12.2%) | 1.097 | 0.739 – 1.629 | 0.953 | 0.631-1.439 |
| Intermediate or above | 107 (24.6%) | 250 (20.0%) | 1.277 | 0.919 – 1.774 | 0.964 | 0.657- 1.416 |
| **Wealth Index** |  |  |  |  |  |  |
| Poorest | 69 (14.4%) | 266 (19.5%) | 1 | 1 | 1 | 1 |
| Poor | 70 (16.8%) | 266 (21.4%) | 1.067 | 0.675 - 1.689 | - | - |
| Middle | 93 (21.8%) | 246 (21.1%) | 1.400 | 0.895 - 2.188 | - | - |
| Rich | 104 (24.7%) | 234 (19.4%) | 1.722* | 1.051 - 2.820 | - | - |
| Richest | 105 (22.3%) | 231 (18.7%) | 1.620 | 0.976 – 2.689 | - | - |
| **Sought ANC** | 408 (92.6%) | 1077 (87.4%) | 1.790* | 1.073 - 2.985 | 1.736* | 1.078-2.796 |
| **4 or more ANC Visits** | 213 (48.8%) | 521 (43.1%) | 1.259* | 0.956 - 1.778 | - | - |
| **FP counseling during ANC Check-up** | 27 (5.9%) | 50 (3.8%) | 1.604* | 0.746 – 3.446 | - | - |
| **LHW Visited during pregnancy** | 304 (69.0%) | 865 (69.2%) | 0.988 | 0.742 - 1.37 | - | - |
| **Facility births** | 361 (81.3%) | 949 (76.5%) | 1.335* | 0.996 - 1.790 | - | - |
| **Skilled Birth Attendant** | 360 (81.1%) | 952 (76.7%) | 1.308* | 0.973- 1.758 | - | - |
| **Type of Delivery** |  |  |  |  |  |  |
| Normal/Assisted | 347 (80.0%) | 1046 (84.9%) | 1 | 1 |  |  |
| C-Section | 94 (20.0%) | 197 (15.1%) | 1.402* | 1.041 - 1.888 | - | - |
| **PNC checkup of mother** | 287 (63.0%) | 707 (56.2%) | 1.330* | 1.008 - 1.754 | 1.466* | 1.093 – 2.052 |
| **LHW Visited after delivery** | 300 (65.7%) | 741 (57.9%) | 1.397* | 1.076 - 1.813 | - | - |
| **LHW counseling for family planning as a part of PNC visit** | 150 (33.6%) | 253 (20.0%) | 2.028* | 1.569 - 2.622 | 1.833* | 1.386- 2.424 |

#### Univariate Analysis

The odds of using contraceptives among women with more than four children were four times higher than those with only one child (OR=3.739; 95% CI=2.274 – 6.149). The likelihood of use of contraceptives was two times higher for women aged 35-39 years compared to women aged below 19 years (OR 2.556 95% CI 1.345-4.857). Women who had completed middle-level education were at least twice as likely to use modern contraceptives to delay pregnancy than those without education (OR 1.629 95% CI 0.970 – 2.737). Similarly, husbands who had completed their middle-level education were at least two times more likely to use contraceptives compared to those who were uneducated (OR 1.879 95% CI 1.201 - 2.939). The odds of modern contraceptive use among women who belonged to the rich wealth index were two times higher than women in the poorest wealth index category (OR = 1.722. 95% CI 1.051 - 2.).

Women who were using health facilities for maternal health services showed a significant increase in the odds of using contraceptives in their postpartum period compared to women who were not using health facilities. Mothers who sought antenatal care services had higher odds of adopting modern contraceptive methods (OR = 1.259 95% CI 0.956-1.778) as compared to those who didn’t have ANC visits. Similarly, mothers who had visited health facilities four or more times for antenatal care checkups were 1.259 times more likely to use contraceptives in their postpartum period than those who had visited less than four times during their pregnancy.

Moreover, mothers who delivered at a health facility showed higher odds of using contraceptives than those who delivered at home (OR = 1.335 95% CI 0.996 - 1.790). The odds of use of contraceptives amongst women with cesarean birth were 1.4 times the odds of use of contraceptives amongst women who had either normal or assisted delivery (95% CI 1.041 - 1.888). Women who had a postnatal checkup also showed higher odds of using postpartum contraceptives than those who didn’t (OR = 1.330 95% CI 1.008 - 1.754). Lady Health Worker visits after delivery and for PNC were associated with a significant increase in the odds of the use of contraception (OR = 2.028 95% CI 1.569 - 2.622). Similarly, family planning counseling during antenatal care visits also showed a significant increase in the odds of the use of postpartum contraceptives compared to those who had not received any counseling (OR = 1.604 95% CI 0.746-3.446).

#### Multivariate Analysis

The multivariate logistic model shows that women with four or more children are four times more likely to use contraceptives than those with only one child (AOR = 5.234 95% CI 2.784 - 9.842). Compared to the bivariate model, after controlling for other covariates, the odds of use of contraceptives were significantly higher among women who had completed primary education compared to uneducated women (AOR = 1.730 95%CI 1.264 – 2.367).The odds ratio remained significant for husbands who had attended middle level education in the multivariate model (AOR = 1.695 95%CI 1.039 - 2.762).The odds of a postnatal checkup of the mother were 1.446 times higher amongst mothers who had used contraceptives (AOR = 1.466 95% CI 1.093-2.052) as well as women who were visited by the Lady Health Worker in their postnatal period and were counseled on family planning (AOR = 1.833 95%CI 1.386- – 2.424).

## Discussion

This study assessed the predictors for opting for voluntary modern contraceptive methods to inform future strategies and decisions. This was achieved through univariate and multivariate analysis for specific variables, including knowledge, practices, access, and use of reproductive and maternal health services. The results showed the number of children, mother’s education, husband’s education, postnatal checkup of mother, and LHW counseling for family planning as part of the PNC visit; were the most identified predictors of modern contraceptive methods. The results provide an insight into the design of family planning and maternal health programs and the required modifications as per the local context.

It was noted that the average number of live births was at least four children per MWRA; women with more than four children were more likely to use modern contraceptives. This showed a significant inclination of the sample population towards limiting their pregnancy rather than spacing between desired pregnancies, particularly in women aged more than 35 years. The majority of them opted for sterilization, a surgical procedure, while younger respondents tend to prefer short-acting methods like condoms, pills, and injections. The utilization of long-acting reversible contraceptives (LARC), such as intrauterine devices (IUDs) and implants, was not widely observed. These patterns among participants align with the results of similar studies conducted in similar contexts. [16, 17, 18].

Conversely, contraception needs related to desire for spacing are predicted to be greater among younger women; however, the results show the reverse whereby contraceptive use amongst younger woman was lower. This dilemma of high need but low use discerned in our results is in line with the findings of previous studies in similar settings [19, 8].

Additionally, the prevalence of contraceptive use among the respondents was almost similar to that reported in the national and regional statistical records [6, 7]. However, the current study has highlighted gaps between the awareness and practice of modern contraceptive methods. Though majority of the participants were aware of contraceptive methods, this did not translate into practice as consistent with other studies [20]. It also highlights the intention to use contraceptives and its translation into practice as based on the willingness motivated by the presence and fulfilment of favorable conditions [21]. This could be indicated as an unmet need for family planning in married couples [22]. Another significant element contributing to narrowing the gap between knowledge and application is the accessibility of contraceptive methods along with their consistent availability. The uninterrupted provision of these methods plays a pivotal role in addressing the issue of unmet needs and fostering the adoption of MCM [23]. Pakistan, as a signatory to the Sustainable Development Goals (SDGs) and a participant in the FP 2020 commitments, has taken a firm stance on ensuring comprehensive access to family planning resources, services, and commodities. Diverse initiatives have been initiated within the country in collaboration with international and domestic non-governmental organizations, with sustained support from National, Provincial, and District governments. A recent development in this context is the introduction of the Costed Implementation Plan (CIP) by the Sindh government under the FP2020-30 initiative [24] which holds the potential to expedite advancements towards achieving the desired mCPR targets while focusing on the management of supply continuity.

Moreover, other negative attitudes and misconceptions, including fear of side effects and a preference for a male baby, continue to contribute to low uptake of contraceptives [22, 25, 8, 18, 26]. Among subgroups in the sample population, the contraceptive prevalence rate was higher in women from the rich quintile, similar to other studies in the local context [8, 27]. Interestingly, women educated at the primary level were more likely to opt for modern contraceptive methods than women who had attained comparatively higher education as opposed to previous studies [27, 28]. Furthermore, in univariate analysis, contraceptive use showed an increase with increasing levels of males’ education. However, this growth was more significant for husbands who attained education until the middle level. These findings are compounded by the literature that describes a comprehensive range of studies on family planning and its association with male engagement [29, 10]. Improving contraceptive uptake and support for opting for methods of choice relies upon the provision of couple counseling and increased male engagement [30].

This study’s most important finding was identifying postnatal checkup (PNC) as the most significant predictor for opting for family planning. Respondents who sought PNC were more likely to use modern contraceptive methods. Evidence in the literature has endorsed that postnatal counseling before discharge significantly improves motivation for and practice of contraceptive uptake [26, 31, 32, 33]. This study also observed that Lady Health Worker’s counselling delivered during a PNC visit increased the likelihood of family planning utilization. The probable cause of this finding can be linked to the preference of the local community for Lady Health Workers to discuss family planning matters on a one-to-one basis at the household level while maintaining confidentiality [18]. Moreover, Lady Health Workers also provide family planning commodities like condoms and pills during their routine visits. Consistent with our results, referral to a health care facility through LHW also increases postpartum utilization of family planning. Therefore, the Lady Health Worker Program strongly influences women’s fertility choices and their use of sexual and reproductive health care services [9].

Our study population reported a lower prevalence of family planning counseling received during ANC visit. At the same time, these visits still influenced positive behaviors towards modern contraceptive methods. In addition to this, family planning exposure was seen to be associated with subsequent antenatal care visits, a higher number of deliveries performed by skilled professionals, and a greater chance of delivery at a health facility [9]. Earlier studies have shown a positive relationship between the delivery of high-quality antenatal family planning counseling services and increased contraceptive use [34, 35].

Another study conducted in a similar setting as our study sites, concluded that in communities where a higher proportion of women received quality antenatal care with an explanation of the importance of postpartum family planning and where discussion of birth spacing was more common, there was significantly higher contraceptive use particularly LARC. [36, 37]. These studies were short-term funded programs highlighting the need for sustainable programs and scaling up existing resources. Thus, multiple determinants then affect contraceptive uptake in the context of Sindh, and strategically addressing these are imperative in improving MCM prevalence rates.

However, what is achievable and sustainable is the expansion of interventions that meet specific needs and target these factors by utilizing existing healthcare delivery systems.

### Limitations and Strengths

A key strength of our cross-sectional study was that we had the opportunity to explore MCM uptake in the context of multiple, complex factors, unique to the context of rural Sindh. This is important, given that rural Sindh is home to highly marginalized and vulnerable communities situated within disaster-prone areas that adversely affect reproductive health outcomes for males and females.

Therefore, this study provides an important evidence-bases that can inform future research, particularly on the qualitative side, to understand these predictors better and further, strategically plan to address them at the community and facility level.

The cross-sectional study design of our study has several limitations on account of self-reported data for multiple factors collected at a single point in time. However, this was controlled by the backward selection of significant indicators for developing a final model of predictors.

Furthermore, we kept sociodemographic factors as predictors for family planning practices, which followed the same pattern as the national trends and were covered by other local studies. The analysis was further expanded for the studied sociodemographic variables to focus mainly on the underlying factors influencing contraception uptake in our study districts. Although the data regarding negative attitudes, concerns about potential side effects, and a preference for male babies was not specifically addressed in the interviews, these factors have been extensively documented as contributing to the low adoption of modern contraceptives.

Another limitation of our study is that our sampled population resided within the catchment area of health facilities and already had some form of access to healthcare services in this regard.

Despite this limitation, our study population’s CPR and MCMs prevalence rate was similar to other regional and national level surveys. This validated the findings that the location of residence, on its own, does not affect contraceptive uptake rates [36], making our sample comparable to other studies.

A further limitation of the cross-sectional nature of this study is that findings cannot be used to analyze all behaviors with regards to contraceptive uptake, given that it has been conducted at one point in time. Understanding predictors requires a more detailed inquiry into motivations, specifically, at the individual level, to adopt MCMs and the risks and challenges faced, in day to day life owing to this. The study tried to mitigate this issue by the use of very detailed questionnaires that comprehensively covered these aspects to a certain extent.

## Conclusion and Recommendations

This study showed that the voluntary uptake of modern contraceptive methods is higher in women receiving skilled-based delivery, postnatal checkups at the facility, and Lady Health Workers visits after delivery. Furthermore, the results showed that women used MCMs for limiting pregnancies rather than spacing and that utilization increases with age. This is an important finding that can be used to inform behavioral change interventions to improve outcomes.

In addition to this, bridging the gap between knowledge and uptake to increase uptake is necessary. Hence, various recommendations have been proposed in the light of this study for providing family planning and maternal, newborn, and child health integrated services through facility-based platforms and community engagement. This is associated with an increased likelihood of modern contraceptive method acceptance. Integrated services must include antenatal and postnatal care visits that incorporate family planning services [38, 31, 39]. Providing these services, calls for the capacity strengthening of Lady Health Workers on family planning counseling regarding method-specific side effects.

A further recommendation of this study is to focus on the introduction of feasible and effective interventions which include in-service training, provision of job aids, and educational materials to health care providers. This will allow them to counsel patients for family planning effectively. At the community level, initiating and consistently monitoring educational sessions or programs highlighting the importance of MCM use to community members is important. It should focus on engaging adolescents through these community-level platforms to educate them about MCM at an appropriate age to gradually reduce social resistance to uptake. A further focus should be on the education of young couples on sexual and reproductive health and MCM. Such education, information, and communication must be developed and delivered within the bounds of privacy and respect for socio-cultural norms. Eventually, the positive impact emanating from these initiatives should be used as an evidence base for advocating for policy changes and reform in reproductive health service delivery and access oriented towards encouraging MCM use and empowering young girls and women to reach their full potential.

## Data Availability

The data will be made avaibale with reasonable request to the corresponding author.

## Acknowledgment

We extend our appreciation to the Department of Health and the Population Welfare Department of the Sindh Government for their essential partnership in project implementation. Our sincere gratitude is directed towards the study participants, whose involvement and input are highly valued. Additionally, we acknowledge the dedicated district-based personnel for their consistent assistance throughout project activities, playing a crucial role in the effective achievement of this initiative.

## Supporting Information

S1_File PDF

